# Ecological Assessment of Transdiagnostic Clinical Symptoms in Serious Mental Illness with Daily Smartphone Surveys

**DOI:** 10.1101/2025.09.26.25336721

**Authors:** Yoonho Chung, Bryce Gillis, Habiballah Rahimi-Eichi, Vincent Holstein, Jeffrey M. Girard, Scott L. Rauch, Dost Öngür, Einat Liebenthal, Justin T. Baker

## Abstract

Clinical symptoms in serious mental illness (SMI) fluctuate dynamically, yet standard interview-based assessments often fail to capture these daily changes. Smartphone-based ecological surveys offer a scalable approach to monitoring symptoms in naturalistic settings. We analyzed longitudinal data from 56 outpatients with psychotic or affective disorders who completed 12,984 daily surveys and 1,028 clinical assessments over one year. Machine learning models showed that smartphone surveys moderately estimated Montgomery–Åsberg Depression Rating Scale (r_rm_ = 0.57; p < 0.001) and Young Mania Rating Scale (r_rm_ = 0.39; p < 0.001) and reliably captured within-person fluctuations. Positive symptoms measured by the Positive and Negative Syndrome Scale were also correlated (r_rm_ = 0.24, p < 0.001), though with variable accuracy across participants. Factor modeling showed strongest convergence in negative affective domains, with symptom severity not affecting adherence. These findings highlight smartphone surveys as an ecologically valid tool for real-time symptom monitoring in SMI.

## Introduction

Expressions of psychopathology in individuals with serious mental illness (SMI), including psychotic and mood disorders, are highly dynamic and vary substantially across individuals.^1,2^ Clinical episodes may emerge rapidly and often lead to significant distress and functional impairment. Tracking this temporal variability is challenging for standard interview-based clinical assessments, as evaluations require trained raters and participants to attend in-person visits, which are costly and logistically burdensome. There is a growing interest for reliable digital phenotyping methods using personal mobile devices that can capture these symptom fluctuations with greater temporal resolution to support the goals of personalized psychiatry.^3,4^ Such tools may offer clinical value by enabling timely clinical decision-making and delivering tailored interventions.^5^ For example, they could help detect early signs of symptom exacerbation or inform the delivery of just-in-time digital mental health support in real-world settings.^6,7^

Currently, structured interviews conducted by clinicians during clinical visits remain the gold standard for psychiatric evaluation. This approach typically relies on retrospective recall of symptoms over weeks or months, which may introduce memory related biases.^8,9^ This is particularly problematic for individuals with SMI, who often experience cognitive impairments.^10,11^ For instance, prior studies have shown that individuals with depressive or psychotic disorders are likely to exaggerate negative affective experiences when recalling them retrospectively, compared to how they rate their emotions in the moment.^9,12^ Individuals with psychosis might show reduced accuracy in self-reflection on their mental health state, further compromising the reliability of retrospective reporting.^13,14^ Furthermore, recalling past experiences in unfamiliar clinical settings may interfere will recall with accuracy due to the difference in pertinent contextual cues.^15^

Ambulatory self-report assessments, including daily diaries and ecological momentary assessment (EMA), offer a way to capture symptoms and experiences as they unfold in everyday life. While EMA traditionally involves multiple within-day prompts to assess momentary states, daily diary surveys represent a lower-frequency but conceptually similar approach, asking participants to reflect on their experiences over the preceding day. Both methods provide intensive longitudinal data that enable researchers to estimate fine-grained symptom trajectories and detect dynamic fluctuations that may be missed by infrequent, retrospective clinical interviews.^16^ By prompting participants to self-report close in time to the experience, ambulatory assessments reduce recall bias and enhance ecological validity.^16,17^ In recent years, the widespread ownership and portability of smartphones have made them an ideal platform for delivering these assessments via push notifications.^18^ 18 Importantly, prior studies show that smartphone-based ambulatory surveys are feasible and acceptable even for individuals with SMI, including those experiencing severe symptoms or low motivation.^19–23^

Despite these advantages, ambulatory assessments also have limitations. They rely on self-report without a clinician to interpret responses within clinical context, raising concerns that cognitive or insight impairments may compromise the accuracy of symptom ratings in SMI.^13,14^ Such factors could introduce bias and inflate variability.^24^ In addition, high-frequency EMA protocols can be burdensome over long periods, sometimes leading to fatigue, burnout, and attrition.^25^ Although evidence is mixed, it is often assumed that cognitive and motivational challenges and overall illness severity may further impede adherence to experience sampling protocols.^23,25–27^ Missing data due to these reasons can substantially reduce the usefulness and interpretability of ambulatory survey resposnes as a symptom monitoring tool.

To evaluate the clinical utility of ambulatory surveys for real-time symptom assessment, two key validation approaches are needed. First, ambulatory survey responses should be examined for convergent validity by testing their correspondence with established interview-based clinical instruments, which (despite their own limitations) remain the gold standard in psychiatry and provide benchmark scores that guide treatment. Second, it is essential to evaluate whether ambulatory surveys could track within-person changes in symptoms over time, not just differences between individuals. A self-report ecological assessment tool must be sensitive to fluctuations in symptoms as they occur, rather than only reflecting stable trait-like differences. To establish this, surveys must be collected over sufficiently long periods to capture episodes of both symptom worsening and improvement within the same individual, allowing tests of whether day-to-day fluctuations in daily ratings reflect meaningful clinical changes rather than random variation or measurement noise.

Prior studies have shown that ambulatory self-report ratings correlate with validated clinical scales in SMI, including schizophrenia, bipolar disorder, and major depressive disorder.^22,28–33^ However, most were brief (a few weeks) and included few clinical assessments, focusing primarily on between-person associations (e.g., group of individuals who report higher average EMA symptoms also have higher average clinical scores). While informative, such cross-sectional effects do not establish that fluctuations in ambulatory self-report ratings covary with temporal changes in clinical symptoms within individuals. Longer study durations spanning multiple clinical assessments are needed to evaluate intra-individual convergence over time.

In this study, we leveraged data from a multi-year digital phenotyping project employing an intensive longitudinal design to track clinically meaningful symptom fluctuations and long-term behavioral trends in individuals with SMI. Participants completed smartphone-based daily diary surveys each evening, reporting on affect, symptoms, and other daily experiences. While EMA traditionally refers to multiple within-day assessments of momentary states, we conceptualize these daily surveys as a lower-frequency form of ambulatory self-report assessment that still enables high-resolution tracking of daily symptom trajectories over the extended period.

Leveraging a transdiagnostic sample, we examined whether machine learning (ML) models could estimate multiple symptom domains assessed with the Positive and Negative Syndrome Scale (PANSS) for psychotic symptoms,^34^ the Young Mania Rating Scale (YMRS) for manic symptoms,^35^ and the Montgomery–Åsberg Depression Rating Scale (MADRS) for depressive symptoms.^36^ We hypothesized that symptom severity estimated from daily surveys would converge with clinician-rated scales, with strongest performance for negative affective domains (depression, anxiety, irritability/hostility) given their direct representation in survey items and higher temporal variability. Finally, we anticipated that survey non-completion would increase with time since enrollment but would not systematically relate to symptom severity or demographics, supporting the feasibility of smartphone-based daily surveys for monitoring within-person trajectories in ecologically valid, scalable ways.

## Materials / Subjects and Methods

### Overview of study design

A total of 56 outpatients with serious mental illness were enrolled in the Bipolar Longitudinal Study between 2015 and 2022, designed to follow participants for up to one year or longer. Participants were recruited through clinician referrals from McLean Hospital’s inpatient units (at the time of discharge) and outpatient psychiatry clinics, including the Schizophrenia and Bipolar Disorder Program. Research staff subsequently contacted referred patients, conducted eligibility screenings, and obtained written informed consent prior to enrollment. Diagnoses included primary psychotic disorders (e.g., schizophrenia, schizoaffective disorder) and affective disorders (e.g., major depressive disorder, bipolar I & II). Participants completed screening, baseline assessments, and monthly follow-ups in person or via video conference call. Trained raters conducted monthly semi-structured interviews using standardized instruments: the PANSS^34^ for psychotic symptoms, YMRS^35^ for manic symptoms, and the MADRS^36^ for depressive symptoms. The study was approved by the Mass General Brigham Institutional Review Board, and all participants provided informed consent.

### Smartphone Survey Data Collection

Participants installed the Beiwe smartphone application to submit daily surveys evaluating daily experiences, including affective states, social activity, stress levels, sleep quality, physical health, and general functioning. Beiwe is an open-source, secure web-based platform designed for digital phenotyping, offering compatibility with both Android and iOS devices. Beiwe offers cloud-based storage compliant with HIPAA standards, and a comprehensive back-end for data processing and analysis.^37^ For this study, the Beiwe was programmed to deliver daily notifications at 5:00 PM Eastern Time, prompting participants to complete a 32-item, and to submit one survey per day.

### Statistical Analyses

#### Smartphone daily survey-derived features

To examine the ability of ecological smartphone survey data to estimate clinical symptom ratings, we applied a two-week time window to extract survey responses preceding each clinical interview. This window was selected to align with standard clinical assessment timeframes and to maximize the overlap between clinical interviews and the availability of smartphone survey data. To assess the robustness of this window, we conducted a sensitivity analysis by varying the duration of the observation window (same day vs 7 vs. 14 days) and the minimum number of completed smartphone surveys required within each window (ranging from 3 to 5 responses). For each clinical assessment that met these inclusion criteria, we derived summary features from the corresponding survey data, including the mean, standard deviation (SD), minimum, and maximum values within the selected time window. These features were used to capture both average levels and variability in symptom expression over time.

#### Machine Learning Model Development and Evaluation

Machine learning model development was conducted using the validated PHOTON AI package.^38^ We evaluated three estimators, elastic net regression, support vector regression (SVR), and random forest regression (RFR), to identify the optimal model for each outcome. Models were trained to minimize mean absolute error (MAE), using participant-level nested leave-one-subject-out cross-validation to prevent data leakage and ensure generalizability. Therefore, when making estimation for each participant with multiple observations, none of the person’s observaions at any point were not used. Hyperparameters were optimized using random grid search with 25 iterations in the inner loops. Optimized hyperparameters were L1 ratio for the elastic net, gamma parameter in the SVR and the number of estimators for the RFR. To assess model significance, p-values were derived from permutation testing comparing observed and estimated values. Feature importance was assessed using permutation feature importance testing with 1000 permutations, to quantify the contribution of each feature to the model’s predictions.

<u>We used repeated measures correlation (rmcorr) to assess the within-subject association between predicted and true values across multiple time points.</u>^39^ Rmcorr estimates the common intra-individual linear association by accounting for non-independence among repeated observations within individuals. Correlation coefficients were interpreted based on conventional thresholds in behavioral and psychological sciences: values between 0.00–0.10 were considered negligible, 0.10–0.39 weak, 0.40–0.69 moderate, 0.70–0.89 strong, and 0.90–1.00 very strong correlations.^40^

To formally account for the repeated-measures structure of the data, we report marginal R² values using mixed-effects regression models. Marginal R² reflects the variance explained by fixed effects (i.e., model predictions). To further characterize variability in model performance at the individual level, we computed within-person Pearson correlations between estimated and observed scores for participants with at least five clinical assessments.

#### Item-level associations

We used Bayesian multilevel modeling approach using the *brms* package in R to examine survey item-level associations between daily survey responses and clinical symptom ratings.^41^ Univariate models were fit separately for each clinical total score using the mean value of each corresponding survey item over a specified time window. Both between-person and within-person effects were estimated simultaneously using standardized slopes. We adopted a univariate modeling framework to enhance interpretability, as feature importance derived from multivariate machine learning models can be difficult to interpret due to multicollinearity and regularization effects.

Between-person effects reflected how individual differences in average survey responses across the study period related to overall symptom severity, captured through individual-level means. Within-person effects assessed whether deviations from an individual’s own average survey ratings were associated with fluctuations in clinical symptoms over time. All models included fixed effects for age and sex, as well as random intercepts and random slopes to account for individual-level variability in baseline symptoms and response trajectories.

Models were estimated using a skew-normal distribution to accommodate potential asymmetry in outcome distributions while preserving positive variance. Weakly informative priors were specified for all parameters, based on the observed mean and standard deviation of each outcome variable. Posterior estimates are summarized using 89% highest density intervals (HDIs), consistent with best practices for Bayesian inference and improved interval stability.^42^ A detailed description of the modeling framework is available in a prior publication,^43^ and model specifications are provided in the supplementary materials.

#### Estimating transdiagnostic symptom dimensions

As a secondary analysis, we trained machine learning models using daily survey data to estimate transdiagnostic symptom factor scores derived from structured clinical assessments. This approach enabled us to assess the extent to which specific latent symptom constructs could be tracked using daily surveys. Latent factor scores were based on a previously validated model that aggregates items from the PANSS, MADRS, and YMRS, developed in a diagnostically heterogeneous sample that closely mirrors the diagnostic composition of the current cohort.^44^ Factor scores were computed for a correlated-factors model comprising seven symptom dimensions: positive symptoms, negative symptoms, depression, mania, disorganization, hostility, and anxiety. Individual-level factor scores were estimated using the Empirical Bayes Modal method, implemented in the *l*avaan R package.^45^

#### Model performance by participant characteristics

To evaluate whether predictive performance varied by diagnosis or sociodemographic characteristics, we compared MAE between participants with a primary psychotic disorder and those with an affective disorder. Primary psychotic disorders included schizophrenia and schizoaffective disorder, while affective disorders included bipolar I disorder, bipolar II disorder, and major depressive disorder, based on DSM-5 diagnoses.^46^ Covariates included age, sex, education level, race, overall EMA missingness, and the mean total scores of each clinical rating scale across all study visits. False discovery rate correction was applied to account for multiple comparisons.

#### Ambulatory survey missingness

Adapting the Bayesian multilevel modeling approach described above, we examined whether daily survey completion rate (i.e., the percentage of daily surveys completed during the 14 days preceding each clinical visit) was associated with each clinical total scores. Number of days since study enrollment, age, and sex were included as covariates.

## Results

### Sociodemographic and BLS data summary

The included sample had a mean age of 30.1 years (SD = 8.8; range = 19–61), and 55.4% identified as female. Consistent with local demographics, the majority of participants were White (64.3%), followed by Asian (16.1%) and African American (14.3%). A total of 89.3% of participants had completed at least some college education. Based on the primary diagnosis, 17 participants were classified with a primary psychotic disorder (i.e., schizophrenia or schizoaffective disorder), and 39 were classified with an affective disorder (i.e., bipolar I disorder, bipolar II disorder, or major depressive disorder).

Duration of study was defined as the period from onboarding to the last clinical assessment with overlapping EMA data that met inclusion criteria in weeks. The median duration of study participation meeting these criteria was 39 weeks (SD = 57). Across the sample, a total of 1,028 interview-based clinical assessments were conducted, with an average of 15.8 (median = 12) clinical interviews per participant (SD = 12.0). A total of 12,984 daily surveys were submitted, with an average of 185.5 daily surveys per participant (SD = 245.0). Sociodemographic and study characteristics stratified by diagnostic group are summarized in Table 1.

**Table 1.**
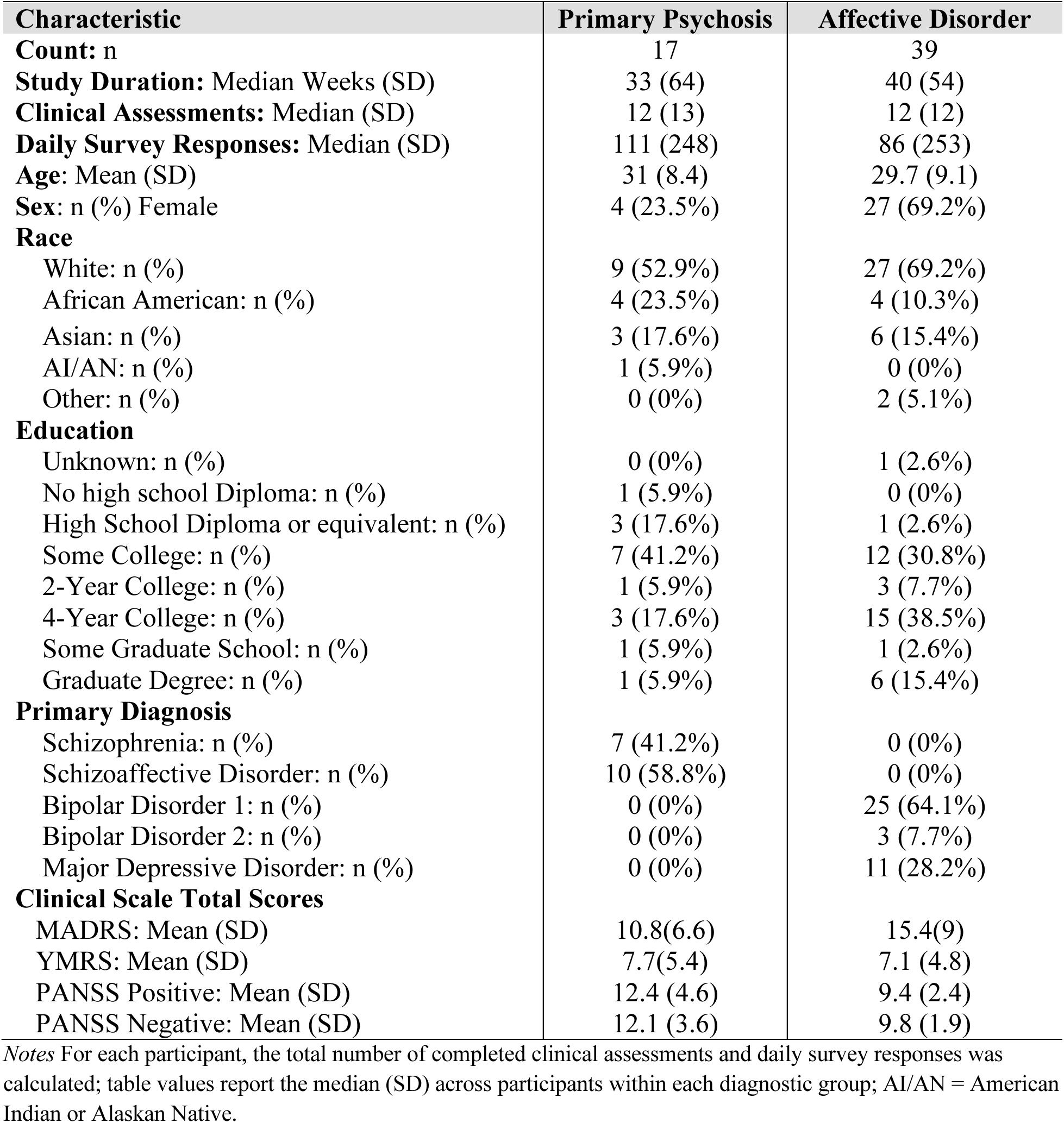
Sociodemographic table of participants with primary psychosis and affective disorder.

### Optimization of observation time windows

Comparisons across daily survey observation windows (same-day, 7-day, and 14-day) and varying survey availability thresholds (3 or 5 daily surveys) indicated that the 14-day window with a minimum threshold of three completed surveys per clinical assessment yielded the highest number of observations (n = 423 instances). The distribution of clinical interview assessments and daily survey responses across different observation window durations and availability thresholds did not reveal statistically significant differences (see Supplementary material).

### Estimating established clinical instruments

Over a 14-day window preceding each clinical interview-based assessment, we extracted summary features (mean, standard deviation, minimum, and maximum) from the ecological data. We evaluated the predictive performance of clinical total scores, including the PANSS Positive, PANSS Negative, MADRS, and YMRS scales. Although overall performance was comparable across models, the elastic net algorithm slightly outperformed SVR and RFR, as indicated by lower MAE. As shown in Figure 1, prediction accuracy was highest for MADRS scores (marginal R² = 0.32; 95% CI [0.25, 0.40]; r_rm_ = 0.57, p < 0.001), indicating moderate predictive performance. Performance for the YMRS was weak (marginal R² = 0.12; 95% CI [0.07, 0.20]; r_rm_ = 0.39, p < 0.001). Prediction of PANSS Positive scores yielded weak performance (marginal R² = 0.04; 95% CI [0.01, 0.07]; r_rm_ = 0.24, p < 0.001), while predictive accuracy for PANSS Negative was negligible (marginal R² = 0.002; 95% CI [0.00, 0.02]; r_rm_ = 0.02, p = 0.66). Linear regression model fits were also stratified by diagnostic group (primary psychosis vs. affective disorder), and no significant group-by-slope interactions were observed. Additional model performance metrics are provided in Table 2 and the Supplementary Materials.

**Figure 1:**
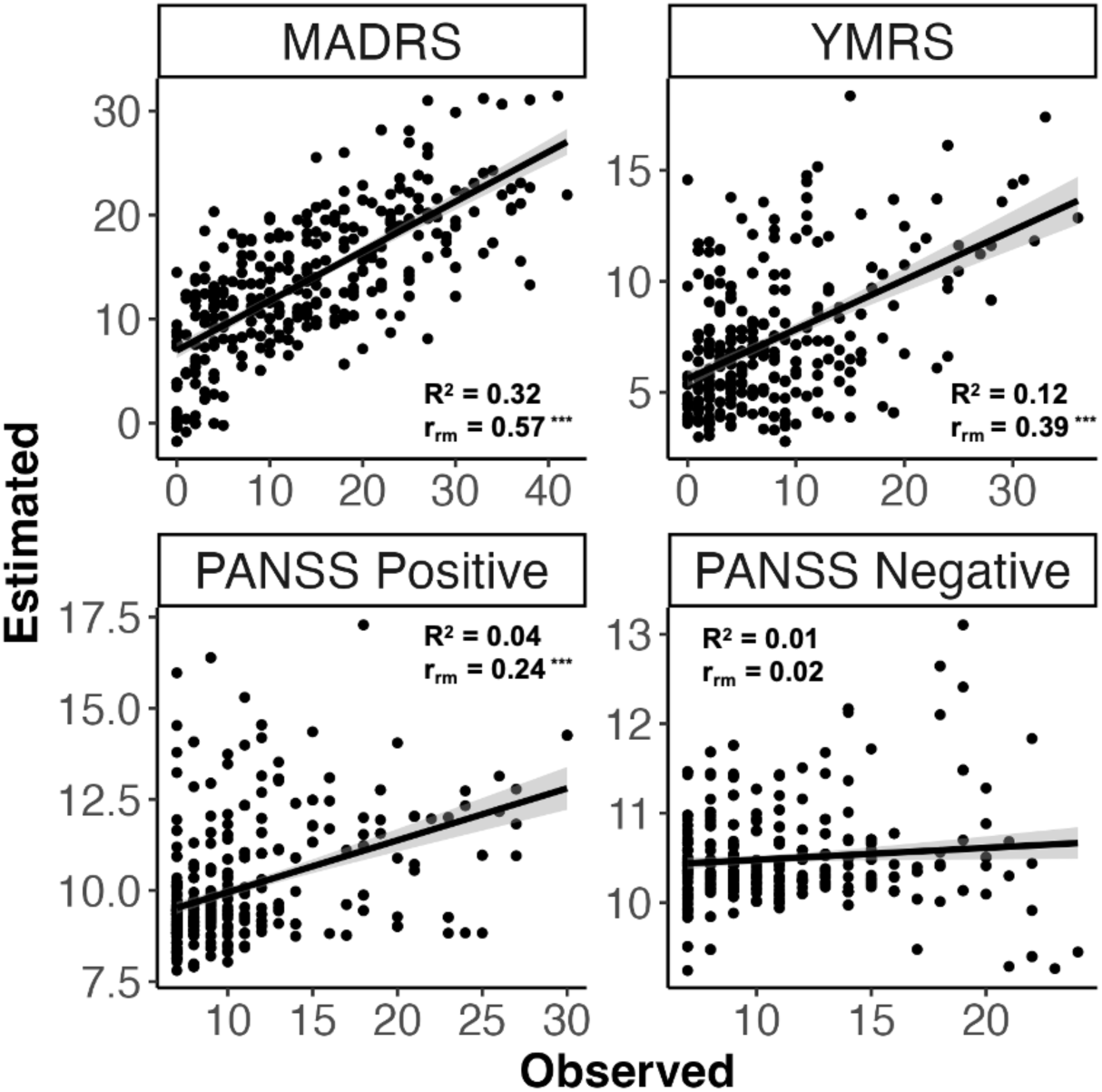
Observed interview-based clinical rating scores plotted against estimates derived from daily smartphone surveys using elastic net regression model.

**Table 2.** Model performance summary for estimating clinical scale scores and factor-based symptom scores.

| Target | MAE | RMSE | $r_{rm}$ | Marginal $R^2$ | Within-Person Corr |
| --- | --- | --- | --- | --- | --- |
| **Clinical Scales** |  |  |  |  |  |
| MADRS | 5.60 | 7.07 | 0.57*** | 0.32 [0.25, 0.40] | 0.48 (0.29) |
| YMRS | 4.60 | 6.00 | 0.39*** | 0.12 [0.07, 0.20] | 0.42 (0.23) |
| PANSS Positive | 2.82 | 4.19 | 0.24*** | 0.04 [0.01, 0.07] | 0.17 (0.33) |
| PANSS Negative | 2.79 | 3.63 | 0.02 | 0.00 [0.00, 0.02] | 0.01 (0.33) |
| **Factor Dimensions** |  |  |  |  |  |
| Depression | 3.37 | 4.22 | 0.43*** | 0.40 [0.29, 0.50] | 0.38 (0.33) |
| Anxiety | 3.84 | 4.74 | 0.41*** | 0.40 [0.27, 0.51] | 0.30 (0.35) |
| Mania | 4.22 | 5.27 | 0.21*** | 0.08 [0.04, 0.18] | 0.05 (0.28) |
| Hostility | 4.03 | 4.86 | 0.32*** | 0.18 [0.09, 0.28] | 0.21 (0.29) |
| Positive Symptoms | 3.40 | 4.76 | 0.31*** | 0.16 [0.06, 0.31] | 0.23 (0.43) |
| Negative Symptoms | 3.79 | 4.85 | 0.23*** | 0.07 [0.02, 0.15] | 0.05 (0.30) |
| Disorganization | 4.05 | 5.14 | 0.19*** | 0.06 [0.01, 0.13] | 0.22 (0.35) |

**Table Note.** *MADRS* = Montgomery–Åsberg Depression Rating Scale; *YMRS* = Young Mania Rating Scale; *PANSS* = Positive and Negative Syndrome Scale; *MAE* = Mean Absolute Error; *RMSE* = Root Mean Squared Error. $R_{rm}$ = Repeated measure correlation. Significance levels: \*\*\* $p < 0.001$ , \*\* $p < 0.01$ , \* $p < 0.05$ . Factor symptoms dimensions are based on Chung et al., 2024. *Within-Person Correlation* represent the mean (SD) of within-person Pearson correlations across individuals.

### Estimating transdiagnostic symptom dimensions

To examine which specific symptom dimensions were most closely tracked by daily survey data, we applied Elastic Net regression models to transdiagnostic factor scores derived from a previously validated dimensional model of serious mental illness. Among all symptom domains, the depression (marginal R² = 0.40; 95% CI [0.29, 0.50]; r_rm_ = 0.43, p < 0.001) and anxiety (marginal R² = 0.40; 95% CI [0.27, 0.51]; r_rm_ = 0.41, p < 0.001) factors demonstrated the highest predictive performance, both in the moderate range. Predictive performance for mania-related dimensions was weaker, with the mania factor showing limited accuracy (marginal R² = 0.08; 95% CI [0.04, 0.18]; r_rm_ = 0.21, p < 0.001) and the hostility factor showing better performance (marginal R² = 0.18; 95% CI [0.09, 0.28]; r_rm_ = 0.32, p < 0.001). Among psychosis-related domains, positive symptoms were predicted at a weak level (marginal R² = 0.16; 95% CI [0.06, 0.31]; r_rm_ = 0.31, p < 0.001), whereas predictive accuracy for negative symptoms (marginal R² = 0.07; 95% CI [0.02, 0.15]; r_rm_ = 0.23, p < 0.001) and disorganization (marginal R² = 0.06; 95% CI [0.01, 0.13]; r_rm_ = 0.19, p < 0.001) were significant, but only accounted for limited variance. Model evaluation metrics for all transdiagnostic symptom dimensions are summarized in Table 2, with additional details provided in the Supplementary Materials.

### Survey item-level associations

As the MADRS demonstrated the most robust predictive performance, we prioritized depressive symptoms in our secondary analyses. Based on Bayesian multi-level regression analyses, survey item-level associations for MADRS, YMRS, PANSS Positive, and PANSS Negative total scores are shown in Figure 3. Higher average ratings of all negative valence items (e.g., anxious, irritable, upset) and feeling “stressed” were significantly associated with greater MADRS score at both the between-person and within-person levels. Conversely, lower ratings of positive valence (e.g., feeling energetic, happy) and lower perceived ability to manage stress were similarly associated with higher MADRS scores at both levels. At the between-person level, greater endorsement of physical or somatic symptoms (e.g., breathing difficulty, heart racing, pain) and lower levels of social engagement (e.g., in-person or digital interaction, feeling outgoing) were also associated with more severe depressive symptoms. Psychosis-related survey items (e.g., auditory or visual hallucinations, paranoid thoughts) were not significantly associated with MADRS scores at either level.

### Within-person associations

Given the robustness of the predictive model for MADRS, we next evaluated its ability to capture temporal variability in MADRS scores at the within-person level. Pearson correlations were calculated within each participant who had at least five clinical assessments (n = 28 out of 56) to characterize individual-level associations between predicted and observed scores. As shown in Figure 2, the distribution of within-person correlations for MADRS scores showed moderate-level associations (median *r* = 0.58, SD = 0.29), suggesting that the machine learning model can significantly estimate MADRS scores over time at the individual level. To further illustrate this, we visualized estimated and observed MADRS scores over time for a subset of participants with more than 10 assessments spanning at least one year, highlighting the range of within-person prediction accuracies. A full summary of within-person correlation results for other clinical scales and factor scores is provided in Table 1 and in the Supplementary Materials.

**Figure 2:**
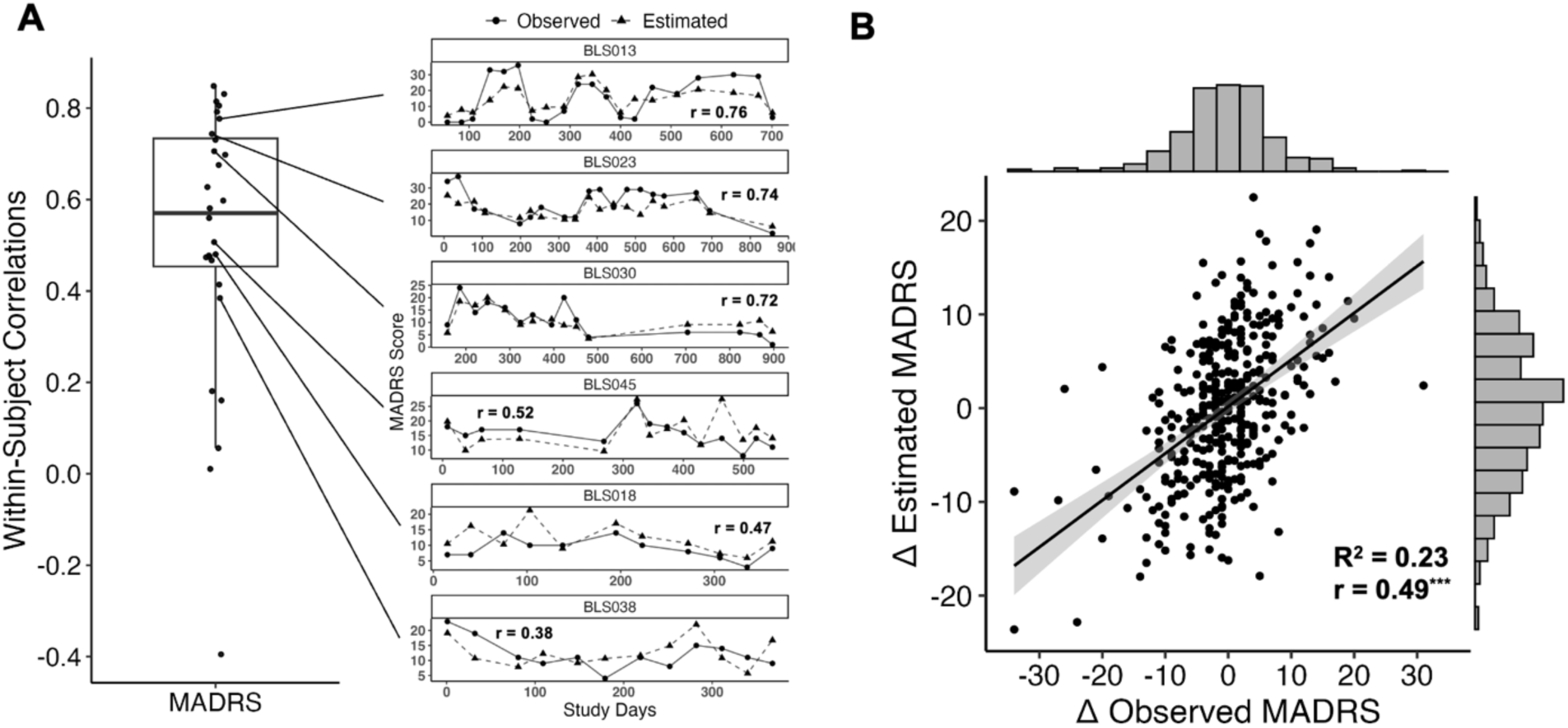
(A) Distribution of within-person correlations between observed and estimated MADRS scores using elastic net, shown as a box plot. Example trajectory scatter plots from six participants with varying degrees of model accuracy are visualized. (B) Estimated change between successive clinical assessments. Estimated change between successive clinical assessments (Δ) derived from daily smartphone surveys plotted against observed changes in clinical rating scores. Marginal distributions are added along each axis.

**Figure 3.**
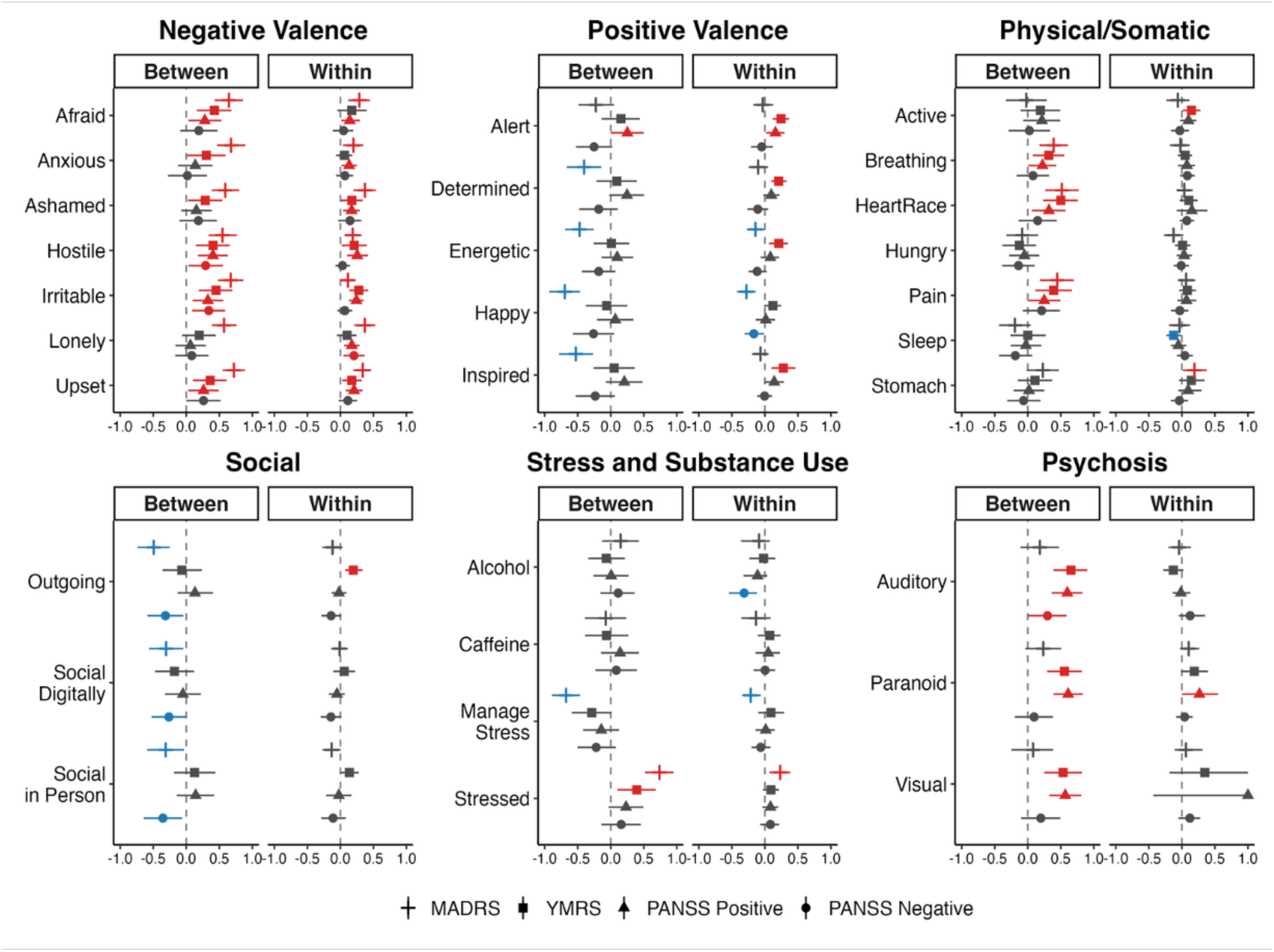
Associations between smartphone-based daily survey items and interview-based clinical scale ratings at the between- and within-person levels using Bayesian multilevel models. Points represent posterior medians, and horizontal bars indicate 89% HDIs. Red indicates significant positive associations; blue indicates significant negative associations.

### Tracking changes in depressive symptoms

We examined whether the model could detect changes in depressive symptoms between subsequent clinical interviews. Change scores (Δ) were computed by calculating the difference in each participant’s observed and predicted MADRS scores at a given visit relative to their immediately preceding visit. As shown in Figure 2B, predicted and observed change scores were moderately correlated (r_rm_ = 0.49, p < 0.001), and the model explained a substantial proportion of the variance in observed change (marginal R² = 0.23).

### Participant characteristics and model performance

No significant differences in MAE were observed between participants with primary psychosis and those with affective disorders across the MADRS, YMRS, PANSS Positive, and PANSS Negative scales (all p > 0.05). Likewise, individual-level predictive performance, assessed by within-person correlations, did not differ significantly between diagnostic groups (all p > 0.05). Other covariates, including age, sex, education, race, overall EMA missingness, and average clinical symptom severity (MADRS, YMRS, PANSS), were not associated with model error (MAE).

### Daily survey missingness

Based on the Bayesian multilevel regression framework described above, higher within-person MADRS scores were associated with greater survey completion rate (14 days prior to each clinical assessment; standardized slope: Median = –0.037, 95% CI [–0.080, –0.001], pd = 0.98). In contrast, between-person variation in MADRS scores was not related to survey completion rate (Median = 0.010, 95% CI [–0.133, 0.163], pd = 0.56). Among covariates, longer study participation was significantly associated with reduced daily survey completion (Median = 0.0003, 95% CI [0.0002, 0.0005], pd = 1.00). Age, sex, and primary diagnosis were not significant predictors. Similarly, within- and between-person effects of YMRS, PANSS Positive, and PANSS Negative scores were all non-significant, with study duration presenting as the only consistent predictor of daily survey missingness across models.

## Discussion

A novel aspect of this study was the use of an intensive longitudinal design in which individuals with SMI, recruited from a transdiagnostic sample, participated in repeated multidimensional clinical assessments while concurrently completing daily smartphone-based surveys over an extended period. This design provided an ecologically valid framework to characterize the natural course of fluctuating symptoms in real-world contexts, addressing a key gap in traditional interview-based assessments. Leveraging this unique dataset, we evaluated whether daily smartphone surveys could reliably estimate face-to-face interview-based symptom severity, with particular attention to the capacity of predictive models to capture dynamic changes at the within-person level.

### Depressive symptoms

Among the clinical symptom scales evaluated, the machine learning model estimating MADRS total scores showed the strongest correspondence with observed clinical ratings. As hypothesized, survey items reflecting negative affect (e.g., feeling lonely, upset, or irritable) and perceived stress were positively associated with MADRS scores at both between- and within-person levels. Notably, the machine learning model estimating depressive symptom scores by the integrated these items, explained significant variance in observed MADRS ratings. Importantly, a change of 6–9 points on the MADRS is considered clinically meaningful, Importantly, a 6–9 point change on the MADRS is considered clinically meaningful, a threshold that exceeded the model’s MAE^47^ In our longitudinal dataset, we identified multiple instances in which participants’ depressive symptoms exceeded this threshold, and we confirmed that the model could track these changes in depressive symptoms at the within-person level, underscoring its potential utility for remote monitoring of changes in depressive symptom severity. These findings underscore the potential utility of this approach for remote monitoring of clinically significant shifts in depressive symptom severity.

A key advantage of our predictive modeling approach is its capacity to generate interpretable depressive symptom severity scores at each survey completion. This allows for daily estimates of symptom severity on a standardized clinical scale, facilitating fine-grained mapping of interpretable symptom trajectories in naturalistic settings without relying on a trained rater. Notably, accuracy of model performance did not differ significantly between individuals with primary psychotic disorders and those with affective disorders, supporting the transdiagnostic applicability of the model.

### Manic Symptoms

Machine learning model for estimating YMRS scores showed moderate performance. Manic experiences often involve comorbid symptoms, including elevated mood, hostility, and irritability, all of which are evaluated with the YMRS. When model accuracy for specific symptom constructs was examined using factor scores, the hostility factor (with strong loadings from hostility and irritability clinical ratings) showed substantially better predictive accuracy than the mania factor (with strong loadings from elevated mood and high energy clinical ratings). This suggests that the model’s performance may be primarily driven by variance related to hostility and irritability. Consistent with this interpretation, daily survey items rating hostility and irritability were significantly associated with YMRS scores at both the between- and within-person levels. In contrast, items reflecting elevated mood (e.g., feeling inspired, determined, or energetic) were associated with YMRS scores only at the within-person level. Similarly, behavioral indicators of mania, such as increased physical activity and reduced sleep, were also associated with YMRS scores at within-person levels. These findings may be attributable to the fact that core manic symptoms such as elevated mood, high energy, and reduced sleep are acute and emerge transiently rather than reflecting stable trait-like differences.

In line with prior work by Busk et al., our findings demonstrate that machine learning models based on smartphone surveys can estimate both depressive and manic symptoms, with substantially greater accuracy for depressive symptoms.^48^ We further expanded prior work by recruiting a transdiagnostic sample, rather than limiting recruitment to individuals with bipolar disorder. Relatively modest predictive performance for YMRS is likely due, at least in part, to the limited number of daily surveys and clinical assessments captured during high manic states. Including additional validated survey items targeting manic symptoms and enrolling individuals with greater symptom severity may help improve model’s performance. However, the underrepresentation of severe manic states may also reflect reduced engagement with ecological sampling methods during acute episodes, as well as the possibility that patients lack insight and under-rate the severity of their symptoms. Further investigation is warranted to clarify these patterns.

### Psychotic Symptoms

The machine learning model estimating PANSS Positive scores was statistically significant but explained only modest variance. Three survey items assessed positive symptoms (visual hallucinations, auditory hallucinations, and paranoid thoughts). Consistent with prior studies, individuals with higher PANSS Positive scores tended to endorse psychosis items with great serverity,^22,31^ but within-person associations were highly variable across participants. Visual inspection of within-person scatter plots revealed that while some individuals showed reasonable correspondence between smartphone survey responses and PANSS Positive ratings over time, many others did not. Notably, several participants with elevated PANSS Positive scores failed to endorse any positive symptom items in their smartphone surveys. This misalignment may reflect reduced insight, or differing interpretations of survey items.^49,50^ Although self-reported survey methods hold promise for capturing positive symptoms in ecological settings, further research is needed to determine for whom this approach provides valid and reliable symptom tracking.

Machine learning models for PANSS Negative symptoms explained negligible variance. This was expected, as the study did not include patients with prominent negative symptoms. Moreover, the survey items included in this study lacked coverage of core negative symptom domains, such as motivation and ability to experience pleasure. The only notable association was at the between-person level, where reduced social engagement (e.g., feeling less socially engaged or outgoing) was linked to higher PANSS Negative scores. Prior studies, however, have shown that EMA items targeting negative symptoms correlate well with validated clinical scales such as the PANSS Negative or Clinical Assessment Interview for Negative Symptoms, and achieved acceptable adherence. ^21,22,51^ Within-person changes, however, have not been systematically investigated, and future studies are needed to evaluate whether these associations hold over time.

### Ecological survey missingness

As expected, study day was a strong predictor of adherence, with response rates to daily survey prompts declining over time. However, increasing symptom severity or any sociodemographic variables were not associated with survey compliance. Contrary to our expectations, participants showed modestly higher compliance rate during periods when their depressive symptoms were elevated relative to their personal average. While further research needs to determine the generalizability of this finding, our results offer encouraging evidence that individuals with SMI can remain engaged with smartphone-based surveys during periods of heightened depressive mood. Future studies should evaluate strategies to further enhance engagement, including incentives that not only promote adherence but also provide direct value to participants, thereby strengthening the utility of EMA in clinical and research contexts.

For other symptom domains, including mania and psychosis, no relationship between survey missingness and symptom severity was observed. However, clinical rating scores in these domains showed limited variability and were heavily skewed toward minimal to moderate symptom levels. Additional observations collected during periods of heightened mania or psychosis will be essential to clarify whether symptom exacerbation influences engagement with smartphone surveys.

### Limitations

Several limitations should be acknowledged. First, the models generally performed poorly in estimating symptoms at the higher end of the severity spectrum, with systematic underestimation for high end values. This pattern is consistent with systematic bias due to do regression-to-the-mean effects commonly observed in regression-based and ML models, which tend to produce conservative estimates at the extremes.^52^ In addition, the limited variability and skewed distribution of clinical ratings toward low symptom severity likely constrained predictive performance, as a restricted symptom range reduces the signal available for model training. Therefore, inclusion of clinical observations during periods of worsened symptoms could significantly enhance model robustness. Second, although we found no evidence that high symptom severity interfered with smartphone survey compliance, our approach could not account for instances in which clinical assessments were missed due to factors such as hospitalization or acute episodes. Consequently, our missingness analysis may not have fully captured patterns of disengagement related to acute clinical episodes. This limitation may have biased the analytic sample toward participants who were more clinically stable and consistently engaged, potentially reducing the generalizability of our findings to individuals experiencing severe symptom fluctuations. Third, the robust models observed for MADRS scores was likely attributable to the relatively large number of survey items assessing experiences of negative motions, which provided a broader pool of relevant predictors. This study did not evaluate the optimal number or specific survey questions needed to estimate symptom severity across different clinical domains. Future research should aim to identify validated items for assessing each symptom domain and to determine the minimal item set required to reach sufficient model accuracy. Such optimization would help reduce participant burden and support sustained long-term engagement.

### Conclusions

Smartphone-based digital health tools offer a scalable approach to monitoring psychiatric symptoms and delivering timely interventions. In this study, we demonstrated that daily ecological surveys could estimate clinical symptom scores on established rating scales without requiring a trained rater in a clinic setting. This approach enables the mapping of dynamic symptom trajectories at high temporal resolution, creating opportunities to detect early signs of symptom worsening and to inform collaborative treatment planning with clinicians. Compared to infrequent face-to-face clinical interviews, which are often impractical due to the burden they place on both participants and clinician resources, ambulatory self-report assessments provide a low-burden means of capturing high-density and interpretable symptom data in real-world settings. Future work should explore integrating these surveys with passive sensor data to further expand monitoring capacity, reduce burden, and increase feasibility for use in routine clinical care.^53,54^

## Data Availability

All data produced in the present study are available upon reasonable request to the authors

## Acknowledgments

YC: This research was supported by the National Institute of Mental Health under Grant No. T32MH016259

